# High prevalence of mixed and non-vaccine HPV oncogenic types and their association with higher mortality in Black women with cervical cancer

**DOI:** 10.1101/2021.01.16.21249558

**Authors:** Rachelle P. Mendoza, Tahmineh Haidary, Elmer Gabutan, Yin Ying Zhou, Zaheer Bukhari, Courtney Connelly, Wen-Ching Lee, Yi-Chun Lee, Raj Wadgaonkar, Raag Agrawal, M.A. Haseeb, Raavi Gupta

## Abstract

We studied the incidence of HPV genotypes in mostly Black women with cervical carcinoma and correlated histopathologic tumor characteristics, immune markers and clinical data with survival. Disease-free survival (DFS) and overall survival (OS) were recorded for 60 months post-diagnosis. Fifty four of the 60 (90%) patients were Black and 36 (60%) were <55 years of age. Of the 40 patients with typeable HPV genotypes, 10 (25%) had 16/18 HPV genotypes, 30 (75%) had one of the non-16/18 HPV genotypes, and 20 (50%) had one of the 7 genotypes (35, 39, 51, 53, 56, 59 and 68) that are not included in the nonavalent vaccine. Mixed HPV infections (≥2 types) were found in 11/40 (27.5%) patients. Patients infected with non-16/18 genotypes, including the most common genotype, HPV 35, had significantly shorter DFS and OS. PD-L1 (p=0.003), MMR expression (p=0.01), clinical stage (p=0.048), histologic grade (p=0.015) and mixed HPV infection (p=0.026) were independent predictors of DFS. A remarkably high proportion of cervical cancer cells in our patients expressed PD-L1 which opens the possibility of the use of immune checkpoint inhibitors to treat these cancers. Exclusion of the common HPV genotypes from the vaccine exacerbates mortality from cervical cancer in underserved Black patients.

---

Cervical cancer is the third most common gynecologic malignancy and cause of death among women in the United States even though a vaccine against HPV, its etiologic agent, has been in use for a decade. Most cervical cancers are squamous cell carcinomas and their epidemiology reveals considerable disparity among races ^1^. The incidence in non-Hispanic blacks is significantly higher (9.5 per 100,000) than that in non-Hispanic whites (1.9 per 100,000) ^1^. Mortality rate is also higher in non-Hispanic blacks (3.8 to 10.1 per 100,000) than that in non-Hispanic whites (2.1 to 4.7 per 100,000) ^1,2^. Five-year survival rates among affected Black women decreased from 64 to 59% between 1974 and 1994, whereas the corresponding survival rates among Caucasian women increased from 70 to 72% ^3^. Disparities in cervical cancer outcomes have been a cause of concern amongst the healthcare community, and has been linked to cultural, socioeconomic and genetic factors ^1^. Studies adjusted for cultural and socioeconomic factors have revealed a biological basis of higher mortality in cervical carcinoma in Black women ^4^. Although the evidence is overwhelming, few studies have focused on Black patients to identify the causes that lead to these disparities. This study was undertaken to explore the HPV genotypes and the biological basis of disparities in cervical carcinoma in women of different racial groups.

Infection with high-risk human papilloma virus (HPV) is one of the definitive events linked to the complex process of cervical carcinogenesis ^1,5^. HPV DNA is identified in 99.7% of cervical cancers, with over 70% being HPV oncogenic types 16 or 18 (16/18) ^1^. Women with cervical tumors infected with HPV types 16/18 have poor survival as compared to those infected with other HPV types. The clinical outcome of infection with HPV types other than HPV 16 and 18 is variable, but large cohort studies have shown better survival and less persistent infection with non-16/18 HPV types ^6^. As new HPV genotypes associated with cervical cancer are identified, their clinical implication is of value in the development of HPV vaccine. In the last 6 years the number of HPV genotypes included in the US Food and Drug Administration (FDA) approved HPV vaccine (Gardasil 9) has increased from 4 to 9, which has the potency to target 70% of the HPV infections. Black patients have a preponderance of non- 16/18 HPV infections, including some rare genotypes with less known pathogenesis and outcomes. More studies on the distribution of HPV genotypes in racially and ethnically diverse patients and a better understanding of pathogenicity of these HPV genotypes are required for a more complete HPV vaccine for better coverage of the vulnerable populations.

The PD-1/PD-L1 axis is one of the immune-checkpoint pathways that have been found to help cancer cells evade the immune response ^7,8^. PD-L1 expression has been studied for determining its therapeutic and prognostic role in cervical cancer. PD-L1 expression in cervical squamous cell carcinoma cells has ranged from 34.4% to 70.1% ^9-11^. PD-L1 overexpression in tumor cells has been found to be associated with lymph node metastasis, vascular invasion, higher International Federation of Gynecology and Obstetrics (FIGO) stage and poor survival ^9,11,12^. PD-1/PD-L1 inhibitors have been approved by the FDA for patients with recurrent or metastatic cancer unresponsive to chemotherapy. However, it can be a potential therapeutic option in patients with early-stage recurrent or locally advanced cervical cancer and who are unresponsive to chemotherapy ^13^. Because of sparse data the efficacy of immune-checkpoint inhibitors has not been established as a first line therapy in primary cervical carcinoma, especially in Black patients, who commonly present with advanced stage disease.

Another biomarker for identifying patients for immunotherapy is the Mismatch Repair (MMR) or Microsatellite Instability (MSI) pathway which was recently approved by the FDA for use in different tumor types including cervical carcinoma ^14,15^. Loss of function of one of the MMR proteins (MLH1, MSH2, MSH6, PMS2) leads to high rates of mutations that accumulate in repetitive nucleotide regions (microsatellites) inactivating the mismatch repair pathway. MMR-deficient tumors have 10-100 times more somatic mutations than MMR-proficient tumors leading to increased tumor antigen burden and immunogenicity. We speculate that MSI may have predictive and prognostic value in cervical cancer along with its co-expression with PD-1/PD-L1 and HPV genotype.

In this study we have attempted to characterize the clinicopathological features of cervical carcinoma in our patients and analyzed the expression of PD-1, PD- L1, MMR and HPV genotyping to explore their pathogenic and therapeutic potential.

## Methods

### Patient Selection

Patients (n = 198) diagnosed with squamous cell carcinoma of the cervix over a 12-year period at a tertiary care hospital were identified. Sixty of these were found to have complete clinical data and adequate histologic specimens, and were grouped based on their race (Black and Other). Histopathological specimens were reexamined to confirm the diagnosis. Paraffin blocks were retrieved, and tissue microarrays were prepared using two 3-mm punches. Clinical data including patient survival were recorded for up to 60 months post-diagnosis. The study was approved by the Institutional Review Board & Privacy Board (IRB) of the State University of New York, Downstate Health Sciences University (IRB #1230970-3).

### Human Papilloma Virus (HPV) Genotyping

Multiplex PCR and agarose gel electrophoresis were performed on DNA extracted from formalin-fixed paraffin-embedded tissue. To evaluate the DNA quantification after DNA extraction, we analyzed DNA measurement using a NanoDrop spectrophotometer (Thermo Fisher Scientific, Waltham, MA). Thirty-three primers designed for high-risk HPV types (16, 18, 26, 31, 33, 35, 39, 45,51, 52, 53, 56, 58, 59, 66, 68, 73, 82) were used, and their unique specificity was confirmed by BLAST analysis(http://www.ncbi.nlm.nih.gov/BLAST/) (Qiagen, Germantown, MD) (Supplementary Table). Primer selection for each reaction tube mix was based on a previous study ^16^. A housekeeping gene (²-globin) common to all high-risk HPV types was utilized as internal control. Specimens that did not yield adequate DNA for HPV typing were retested. PCR products were analyzed by electrophoresis on a 2% agarose gel stained with ethidium bromide, band sizes were estimated by comparison with a 100 bp molecular weight marker (Invitrogen 100 bp DNA Ladder), and gels were photographed in a UV transilluminator (Thermo Fisher Scientific, Waltham, MA) (Fig. 1).

**Figure 1.**
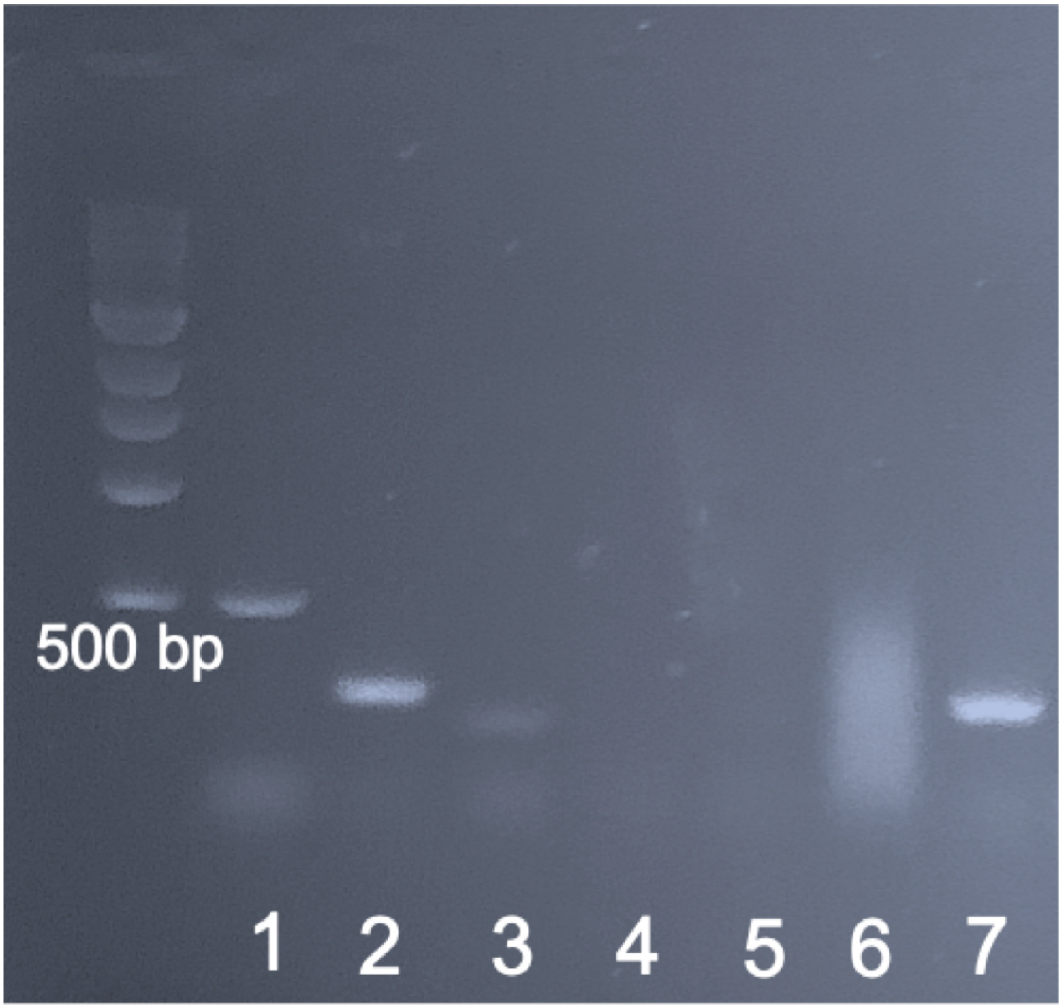
A: A representative agarose gel of HPV PCR products from one cervical cancer specimen with HPV 18. Lane 1: HPV 18 detectable as 536 bp band; Lane 2: HPV type 18 detectable as 274 bp band; Lanes 3-6: No HPV detected; Lane 7: β-globin (positive control). See Supplementary Table for a complete list of primers used.

### Immunophenotyping

PD-L1 (SP263), PD-1, MMR proteins (MSH2, MSH6, PMS2 and MLH1) and p16 expression was characterized by immunohistochemistry (Ventana BenchMark Ultra, Oro Valley, AZ) following the manufacturer’s protocol.

At least 100 viable tumor cells were evaluated for each of the proteins. PD-L1 expressing tumor cells (moderate to strong membranous staining) were quantified as negative (<1% positive cells), low expression (>1 to 49% positive cells), or high expression (>50% positive cells) (Fig. 2). Strong membranous PD- 1 expression in tumor-infiltrating lymphocytes (TILs) was categorized as low or high based on values below or above, the 50th percentile of PD-1+ TILs/mm^2^, respectively. MMR proteins were evaluated by the presence or absence of nuclear staining in the tumor cells. An “intact” status was assigned to specimens with unequivocal nuclear staining in viable tumor cells. “Loss” of expression was assigned to complete loss of nuclear staining or focal weak or equivocal staining in the viable tumor cells in the presence of internal positive controls (lymphocytes, fibroblasts or normal epithelium) ^17^. Immuno- histochemical expression for p16 was defined as “positive” if there was diffuse strong nuclear positivity in tumor cells, while “negative” included those with patchy, wild-type p16 staining pattern ^18^.

**Figure 2.**
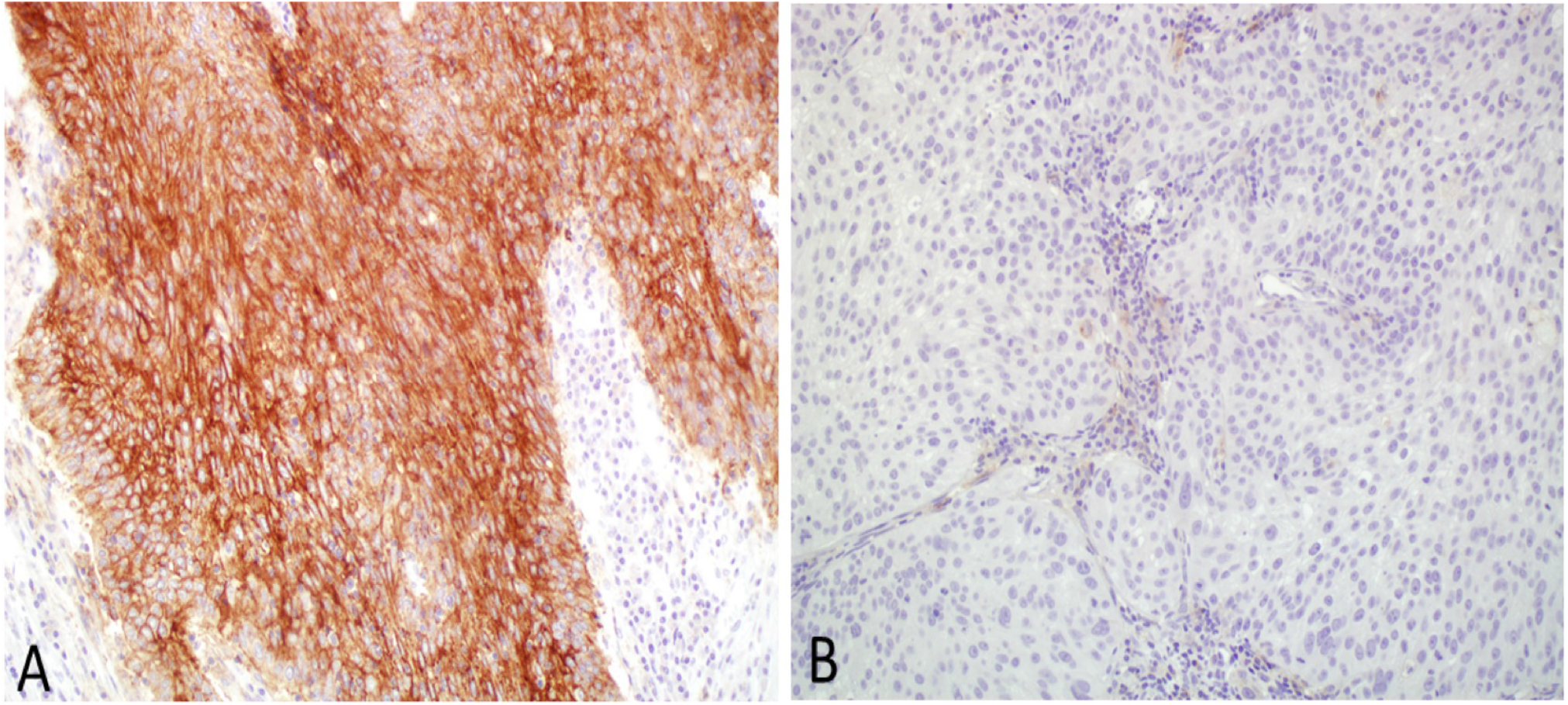
Cervical tumors with PD-L1 expression (A), and without PD-L1 expression (B). x1000.

### Statistical Analysis

Age was categorized using 55 years as cut-off for childbearing age. Clinical staging was determined after surgical management of cases and was based on International Federation of Gynecology and Obstetrics (FIGO) 2009 classification. Clinical stages I and II were combined into “early” clinical stage and stages III and IV as “late” clinical stage. Histologic grades were based on the extent of cellular differentiation, and categorized into well, moderate and poor. Patients were followed for up to 60 months after initial diagnosis. Disease-free survival (DFS) was computed based on the number of months from diagnosis to recurrence, and overall survival (OS) was the number of months from diagnosis to death. Cases with incomplete follow up were censored.

The results of the HPV PCR analysis were recorded as positive or negative (Fig. 3). Nine cases that tested positive only for the common high-risk HPV â-globin gene were treated as a separate group. Cases with specific type results were further subcategorized based on HPV genotypes [16 or 18 (16/18)], non-16/18, and mixed 16/18+ non-16/18), mixed HPV infections (1 or >1 HPV genotype) and based on vaccine coverage (vaccine covered and nonvaccine covered HPV). Most common HPV genotypes (16, 18 and 35) were analyzed against all other HPV genotypes. When examining a specific HPV genotype, it includes all patients with that HPV genotype (including single and mixed infections). Patients categorized as “vaccine covered” had positive PCR results for HPV high-risk genotypes covered by the currently FDA-approved nonavalent vaccine (Gardasil 9), which includes high-risk types 16, 18, 31, 33, 45, 52, and 58.

**Figure 3.**
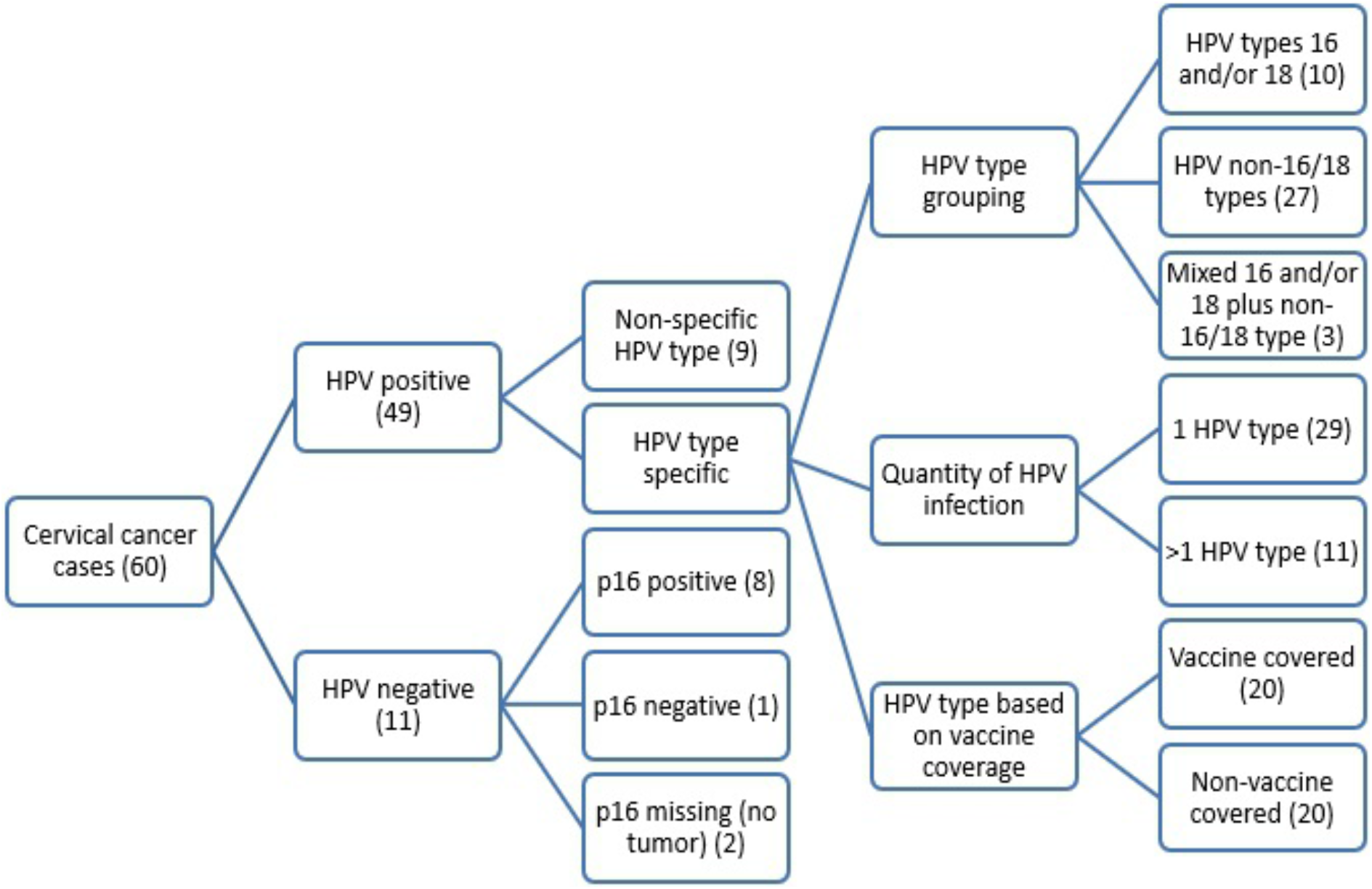
Schematic representation of patient groups relative to HPV genotypes.

Correlations between different parameters were determined by Pearson Chi-Square or Fisher’s exact test. All reported p values are two sided. A p < 0.05 was considered significant. Cumulative 60-month survival was calculated by the Kaplan-Meier method and analyzed by log rank test (SPSS, ver. 26).

## Results

### Patient Characteristics

Sixty patients diagnosed with squamous cell carcinoma of the cervix were included in this study. All of the patients received standard treatment protocol as per the National Comprehensive Cancer Network (NCCN) guidelines based on clinical stage (FIGO classification). Treatment ranged from loop electrosurgical excision to radical hysterectomy with lymph node dissection. The mean age at diagnosis was 52.8 (range: 27 - 86) years. 54 (90%) patients were Black and 6 (10%) were Other (3 white; 3 others). 44 (73.3%) patients were diagnosed at an early clinical stage (I & II). About half of the patients’ tumor histologic grade was poor (51.7%), followed by moderate (36.7%) and well (11.7%) (Table1).

**Table 1.** Demographic and clinical features of patients with squamous cell carcinoma of the cervix.

| **Features** | **Categories** | **n (%)** |
| --- | --- | --- |
| **Age** | <55 years | 36 (60) |
|  | ≥ 55 years | 24 (40) |
| **Race** | Black | 54 (90) |
|  | Other | 6 (10) |
| **Clinical stage** | Early | 44 (73.3) |
|  | Late | 16 (26.7) |
| **Histologic grade** | Well differentiated | 7 (11.7) |
|  | Moderately differentiated | 22 (36.7) |
|  | Poorly differentiated | 31 (51.7) |
| **Outcome** | Alive | 36 (60) |
|  | Alive with recurrence | 4 (6.7) |
|  | Deceased | 14 (23.3) |
|  | Lost to follow up | 6 (10) |
| **HPV PCR result** | Negative | 11 (18.3) |
|  | Positive | 49 (81.7) |
| **HPV type grouping** | Types 16 and/or 18 | 10 (16.7) |
|  | Types non-16 and non-18 | 27 (45) |
|  | Mixed 16/18 and non-16/18 | 3 (5) |
|  | Nontypeable* | 9 (15) |
|  | Negative | 11 (18.3) |
| **Quantity of HPV infection** | 1 HPV type | 29 (72.5) |
|  | >1 HPV type | 11 (27.5) |
| **HPV type based on vaccine coverage\*\*** | Vaccine covered HPV types | 20 (50.0) |
|  | Non-vaccine covered HPV type(s) | 20 (50.0) |

\*Nontypeable HPV genotypes are positive only for the common high-risk HPV β-globin gene.
\*\*Vaccine covered high-risk HPV genotypes are included in the FDA-approved nonavalent vaccine, Gardasil (16, 18, 31, 33, 45, 52 and 58).

### Human Papilloma Virus (HPV) Genotyping

HPV DNA analysis yielded results in 49 of 60 (81.7%) patients; 9 patients had nontypeable HPV genotypes. Thirteen high-risk HPV types (16, 18, 33,35, 39, 45, 51, 52, 53, 56,58, 59, 68) were identified. HPV non-16/18 were present in 27 of 60 (45%) patients, HPV 16/18 in 10 (16.7%) and 3 (5%)patients had both types 16/18 and non-16/18 HPV (types 35, 45 and 52). The most frequently isolated HPV types were 35 (8/40; 20%), followed by 18 and 16 (7/40, 17.5% each). Mixed HPV infections (>1 type) were found in 11 of 40 (27.5%) patients, and all mixed cases had at least one non-16/18 HPV type. None of the patients had both HPV 16 and 18. HPV type 58 was detected exclusively in mixed infections (5/11). Twenty of 40 (50%) of the patients were infected with at least 1 non-vaccine HPV genotype, which included 35, 39, 51, 53, 56, 59 and 68 (Fig. 4). Nine of 60 (15%) patients who had nontypeable HPV genotypes tested positive for viral â-globin but no specific genotype was detected by the 35 primers used.

**Figure 4.**
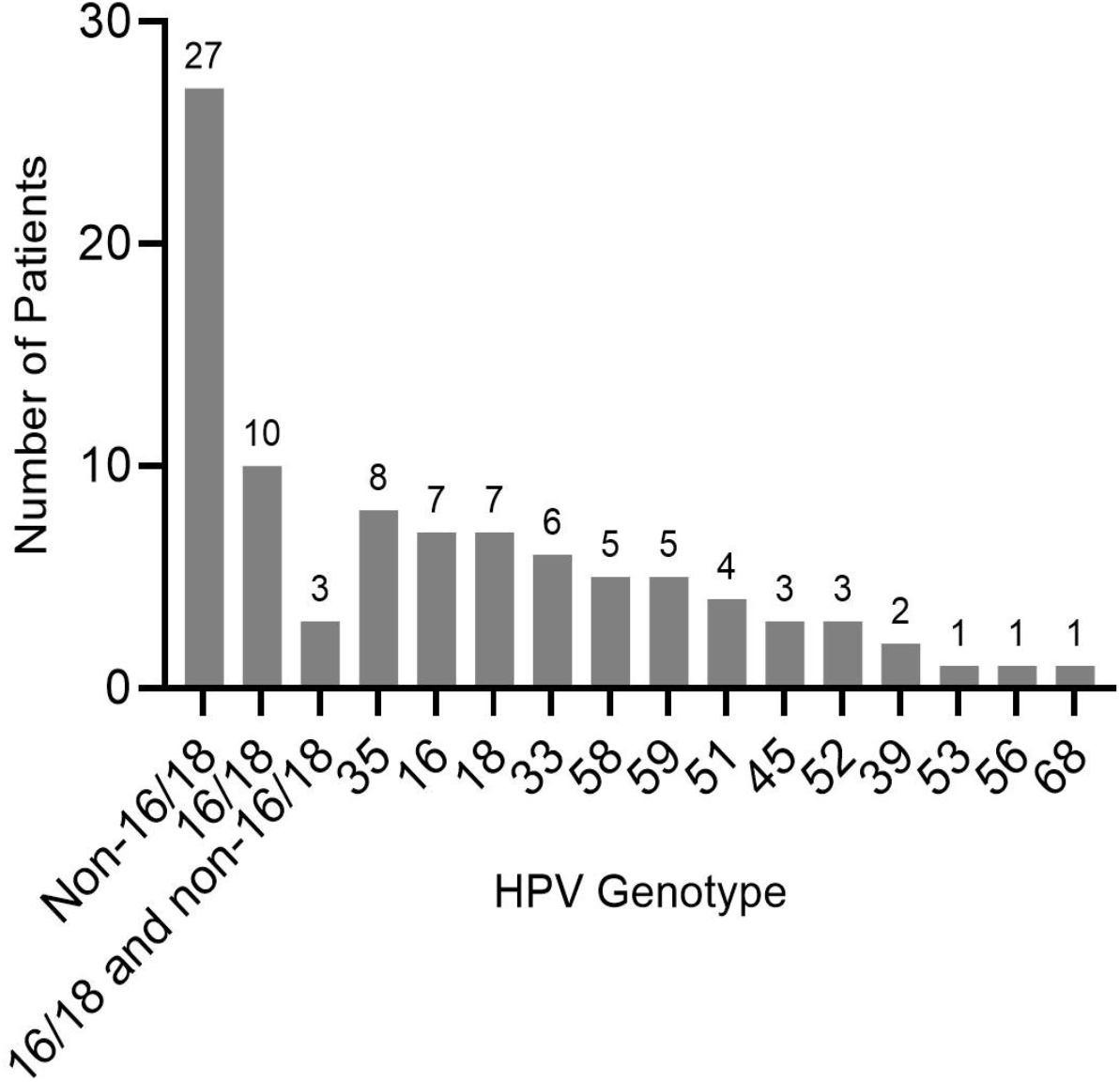
Frequency of HPV types isolated from squamous cell carcinoma of the cervix.

### Immunophenotyping

PD-L1 expression was observed in 56 of 60 (93.3%) patients with 34 (56.7%) showing high and 22 (36.7%) showing low expression. PD-1 expression in TILs was high in 33 of 60 (55%) and low in 27 (45%) patients. MMR was intact in 55 of 60 (91.7%) and lost in 5 (8.3%) patients (2 lost MSH2/MSH6, 1 lost MLH1/PMS2 and 2 lost PMS2). Tissue was lost for 5 patients during immunohistochemical analysis for p16; it was positive in 50 of 55 (90.9%) patients, and negative in 5 (9.1%). Nine out of the 11 patients with negative HPV PCR results had adequate amount of tumor tissue for p16 analysis; 8 of them had positive p16 expression and 1 was negative for p16 (Fig. 3).

### Correlation Analysis

HPV genotypes 16/18 were present in 13/40 (25%) patients in the study. Most (70%) of these patients were <55 years of age and 90% were diagnosed at an early clinical stage with poor histologic tumor grade (6/10; 60%). Patients with non-16/18 HPV genotypes were present in 27/40 (67.5%) patients. They presented later in life (>55 years), mostly diagnosed at an early clinical stage 21/27 (77.8%), and 15/27 (56%) had well to moderate histologic grade tumor. Both groups have a similar recurrence (30%) and death rate (20%). It is noteworthy that, although the non-16/18 HPV tumors had a lower histological grade, patients had a poor prognosis.

Poor histologic grade was most commonly seen in patients with HPV 18 (6/7; 85.5%; p=0.045) followed by HPV genotype 35 (5/8; 62.5%; p=0.694), non- 16/18 HPV (12/27; 44%; p=0.379) and HPV 16 (3/7; 42%; p=0.364) (Table 2).HPV 35 infection was seen predominantly in Black patients (7/8; 88%; p=0.498). It presents in younger patients with a high rate of recurrence. Higher proportion (63.5%) of HPV 35 patients had poor tumor histologic grade as compared to other genotypes (46.9%) (p=0.694). The recurrence and death rates of patients with HPV 35 were higher than those with other HPV genotypes (50% vs 34.5%, and 33.3% vs 27.6%, respectively).

**Table 2.**
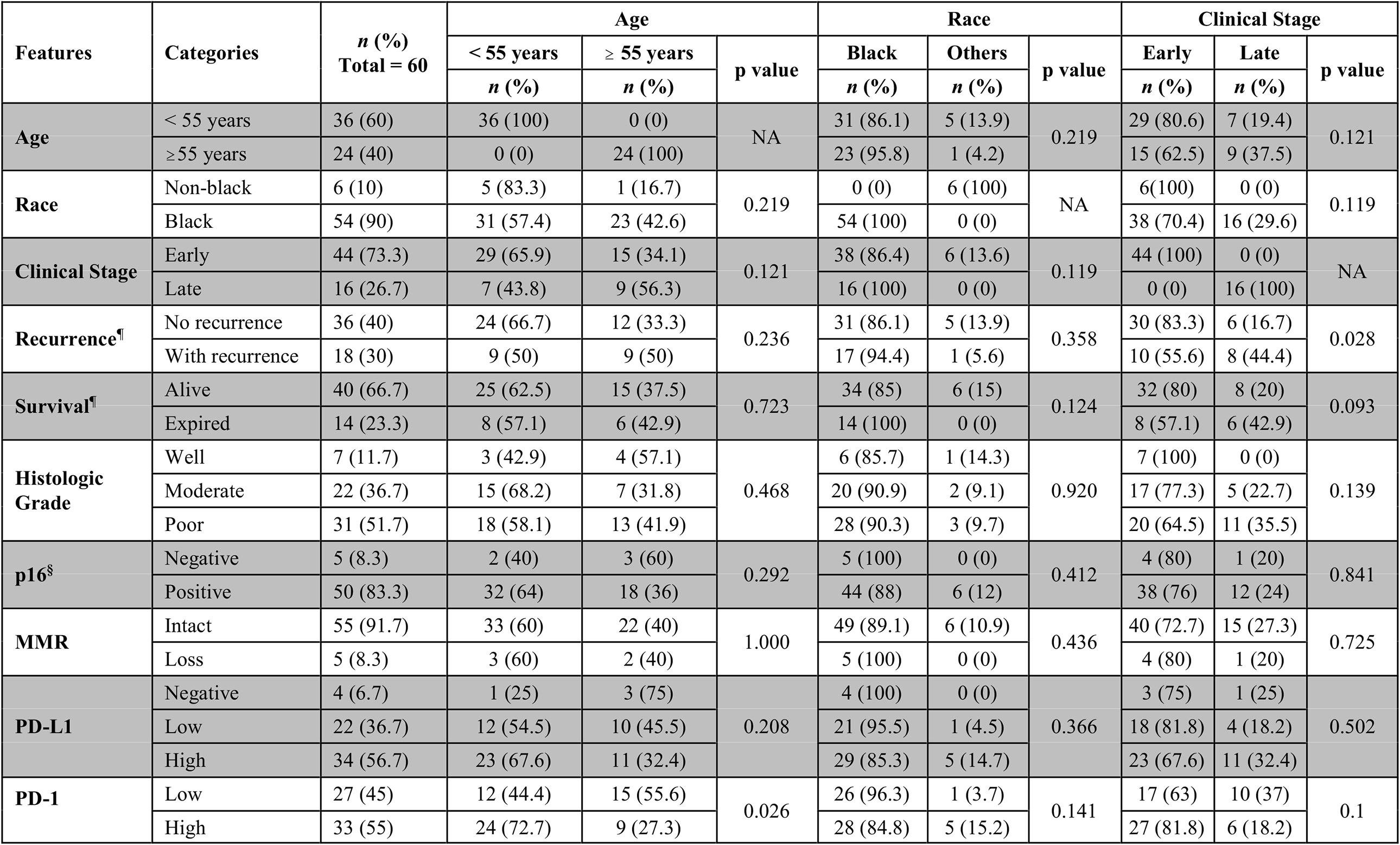
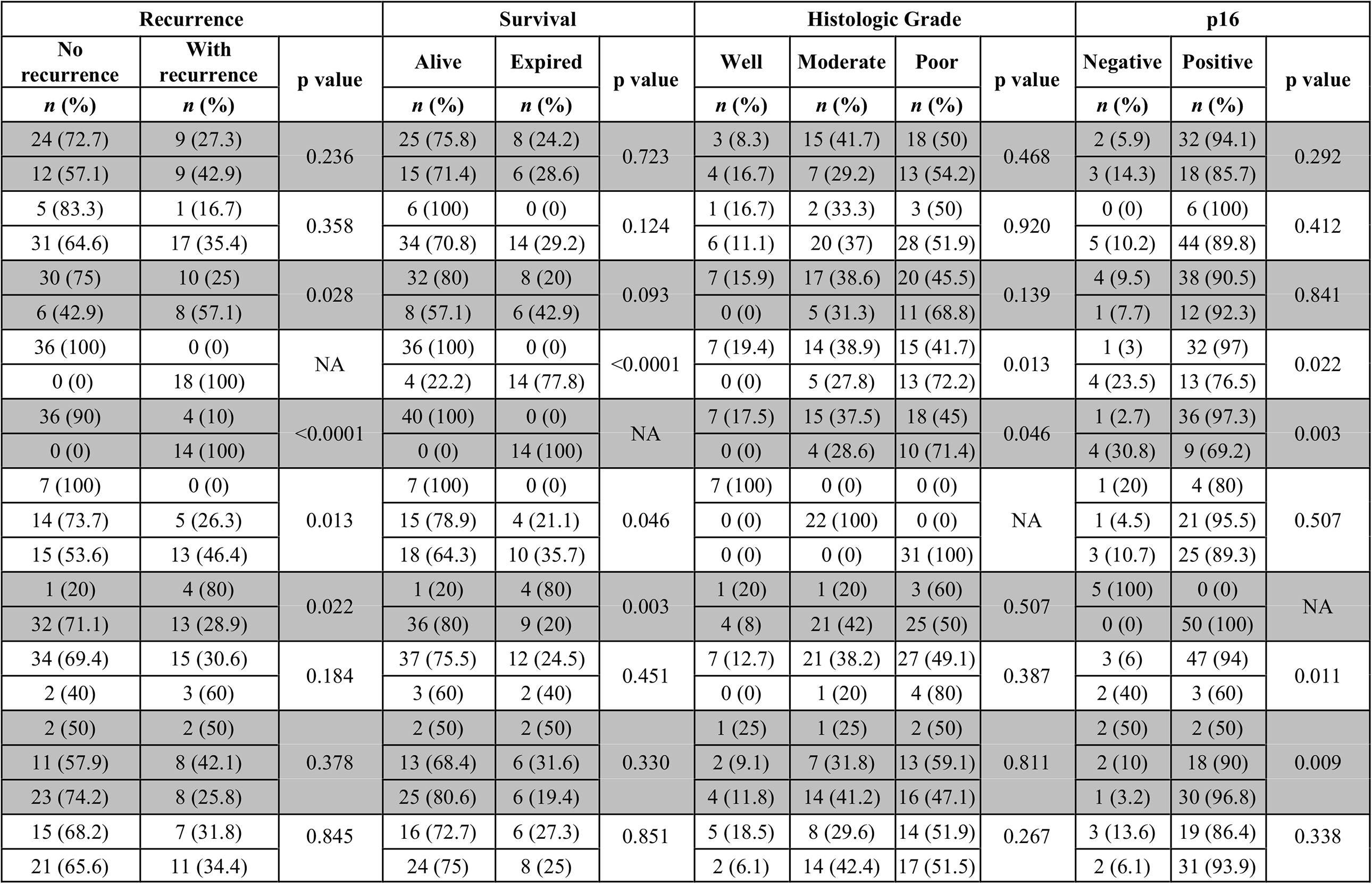
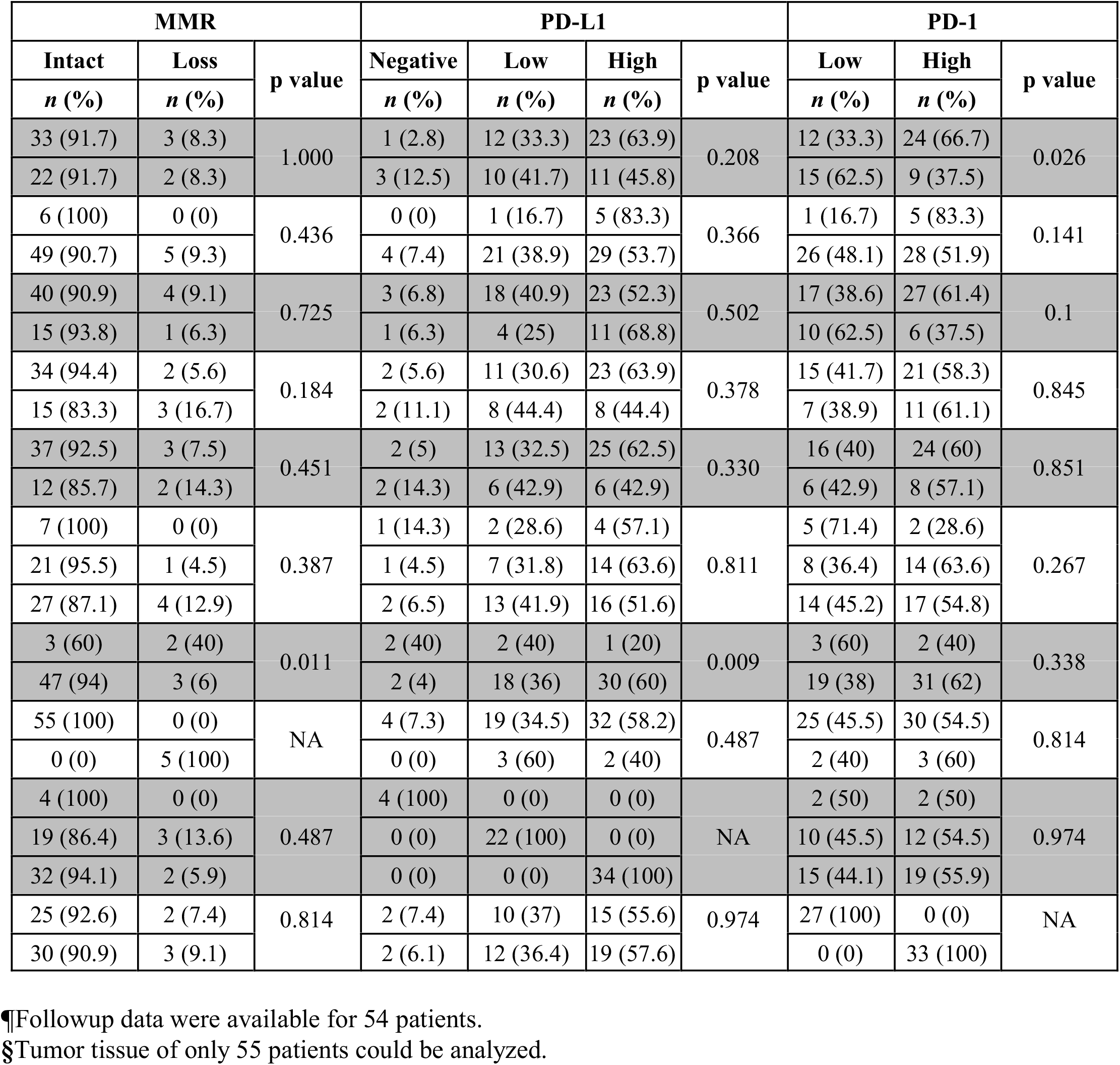
Clinicopathologic features of patients in relation to p16, MMR status, PD-L1 and PD-1 expression in tumor infiltrating lymphocytes.

| Features | Categories | n (%)<br>Total = 60 | Age |  | p value | Race |  | p value | Clinical Stage |  |  |
| --- | --- | --- | --- | --- | --- | --- | --- | --- | --- | --- | --- |
|  |  |  | < 55 years | ≥ 55 years |  | Black | Others |  | Early | Late | p value |
|  |  |  | n (%) | n (%) |  | n (%) | n (%) |  | n (%) | n (%) |  |
| Age | < 55 years | 36 (60) | 36 (100) | 0 (0) | NA | 31 (86.1) | 5 (13.9) | 0.219 | 29 (80.6) | 7 (19.4) | 0.121 |
|  | ≥ 55 years | 24 (40) | 0 (0) | 24 (100) |  | 23 (95.8) | 1 (4.2) |  | 15 (62.5) | 9 (37.5) |  |
| Race | Non-black | 6 (10) | 5 (83.3) | 1 (16.7) | 0.219 | 0 (0) | 6 (100) | NA | 6 (100) | 0 (0) | 0.119 |
|  | Black | 54 (90) | 31 (57.4) | 23 (42.6) |  | 54 (100) | 0 (0) |  | 38 (70.4) | 16 (29.6) |  |
| Clinical Stage | Early | 44 (73.3) | 29 (65.9) | 15 (34.1) | 0.121 | 38 (86.4) | 6 (13.6) | 0.119 | 44 (100) | 0 (0) | NA |
|  | Late | 16 (26.7) | 7 (43.8) | 9 (56.3) |  | 16 (100) | 0 (0) |  | 0 (0) | 16 (100) |  |
| Recurrence<sup>¶</sup> | No recurrence | 36 (40) | 24 (66.7) | 12 (33.3) | 0.236 | 31 (86.1) | 5 (13.9) | 0.358 | 30 (83.3) | 6 (16.7) | 0.028 |
|  | With recurrence | 18 (30) | 9 (50) | 9 (50) |  | 17 (94.4) | 1 (5.6) |  | 10 (55.6) | 8 (44.4) |  |
| Survival<sup>¶</sup> | Alive | 40 (66.7) | 25 (62.5) | 15 (37.5) | 0.723 | 34 (85) | 6 (15) | 0.124 | 32 (80) | 8 (20) | 0.093 |
|  | Expired | 14 (23.3) | 8 (57.1) | 6 (42.9) |  | 14 (100) | 0 (0) |  | 8 (57.1) | 6 (42.9) |  |
| Histologic Grade | Well | 7 (11.7) | 3 (42.9) | 4 (57.1) | 0.468 | 6 (85.7) | 1 (14.3) | 0.920 | 7 (100) | 0 (0) | 0.139 |
|  | Moderate | 22 (36.7) | 15 (68.2) | 7 (31.8) |  | 20 (90.9) | 2 (9.1) |  | 17 (77.3) | 5 (22.7) |  |
|  | Poor | 31 (51.7) | 18 (58.1) | 13 (41.9) |  | 28 (90.3) | 3 (9.7) |  | 20 (64.5) | 11 (35.5) |  |
| p16<sup>§</sup> | Negative | 5 (8.3) | 2 (40) | 3 (60) | 0.292 | 5 (100) | 0 (0) | 0.412 | 4 (80) | 1 (20) | 0.841 |
|  | Positive | 50 (83.3) | 32 (64) | 18 (36) |  | 44 (88) | 6 (12) |  | 38 (76) | 12 (24) |  |
| MMR | Intact | 55 (91.7) | 33 (60) | 22 (40) | 1.000 | 49 (89.1) | 6 (10.9) | 0.436 | 40 (72.7) | 15 (27.3) | 0.725 |
|  | Loss | 5 (8.3) | 3 (60) | 2 (40) |  | 5 (100) | 0 (0) |  | 4 (80) | 1 (20) |  |
| PD-L1 | Negative | 4 (6.7) | 1 (25) | 3 (75) | 0.208 | 4 (100) | 0 (0) | 0.366 | 3 (75) | 1 (25) | 0.502 |
|  | Low | 22 (36.7) | 12 (54.5) | 10 (45.5) |  | 21 (95.5) | 1 (4.5) |  | 18 (81.8) | 4 (18.2) |  |
|  | High | 34 (56.7) | 23 (67.6) | 11 (32.4) |  | 29 (85.3) | 5 (14.7) |  | 23 (67.6) | 11 (32.4) |  |
| PD-1 | Low | 27 (45) | 12 (44.4) | 15 (55.6) | 0.026 | 26 (96.3) | 1 (3.7) | 0.141 | 17 (63) | 10 (37) | 0.1 |
|  | High | 33 (55) | 24 (72.7) | 9 (27.3) |  | 28 (84.8) | 5 (15.2) |  | 27 (81.8) | 6 (18.2) |  |

| Recurrence |  |  | Survival |  |  | Histologic Grade |  |  | p16 |  |  |  |
| --- | --- | --- | --- | --- | --- | --- | --- | --- | --- | --- | --- | --- |
| No recurrence | With recurrence | p value | Alive | Expired | p value | Well | Moderate | Poor | p value | Negative | Positive | p value |
| *n* (%) | *n* (%) |  | *n* (%) | *n* (%) |  | *n* (%) | *n* (%) | *n* (%) |  | *n* (%) | *n* (%) |  |
| 24 (72.7) | 9 (27.3) | 0.236 | 25 (75.8) | 8 (24.2) | 0.723 | 3 (8.3) | 15 (41.7) | 18 (50) | 0.468 | 2 (5.9) | 32 (94.1) | 0.292 |
| 12 (57.1) | 9 (42.9) |  | 15 (71.4) | 6 (28.6) |  | 4 (16.7) | 7 (29.2) | 13 (54.2) |  | 3 (14.3) | 18 (85.7) |  |
| 5 (83.3) | 1 (16.7) | 0.358 | 6 (100) | 0 (0) | 0.124 | 1 (16.7) | 2 (33.3) | 3 (50) | 0.920 | 0 (0) | 6 (100) | 0.412 |
| 31 (64.6) | 17 (35.4) |  | 34 (70.8) | 14 (29.2) |  | 6 (11.1) | 20 (37) | 28 (51.9) |  | 5 (10.2) | 44 (89.8) |  |
| 30 (75) | 10 (25) | 0.028 | 32 (80) | 8 (20) | 0.093 | 7 (15.9) | 17 (38.6) | 20 (45.5) | 0.139 | 4 (9.5) | 38 (90.5) | 0.841 |
| 6 (42.9) | 8 (57.1) |  | 8 (57.1) | 6 (42.9) |  | 0 (0) | 5 (31.3) | 11 (68.8) |  | 1 (7.7) | 12 (92.3) |  |
| 36 (100) | 0 (0) | NA | 36 (100) | 0 (0) | <0.0001 | 7 (19.4) | 14 (38.9) | 15 (41.7) | 0.013 | 1 (3) | 32 (97) | 0.022 |
| 0 (0) | 18 (100) |  | 4 (22.2) | 14 (77.8) |  | 0 (0) | 5 (27.8) | 13 (72.2) |  | 4 (23.5) | 13 (76.5) |  |
| 36 (90) | 4 (10) | <0.0001 | 40 (100) | 0 (0) | NA | 7 (17.5) | 15 (37.5) | 18 (45) | 0.046 | 1 (2.7) | 36 (97.3) | 0.003 |
| 0 (0) | 14 (100) |  | 0 (0) | 14 (100) |  | 0 (0) | 4 (28.6) | 10 (71.4) |  | 4 (30.8) | 9 (69.2) |  |
| 7 (100) | 0 (0) | 0.013 | 7 (100) | 0 (0) | 0.046 | 7 (100) | 0 (0) | 0 (0) | NA | 1 (20) | 4 (80) | 0.507 |
| 14 (73.7) | 5 (26.3) |  | 15 (78.9) | 4 (21.1) |  | 0 (0) | 22 (100) | 0 (0) |  | 1 (4.5) | 21 (95.5) |  |
| 15 (53.6) | 13 (46.4) |  | 18 (64.3) | 10 (35.7) |  | 0 (0) | 0 (0) | 31 (100) |  | 3 (10.7) | 25 (89.3) |  |
| 1 (20) | 4 (80) | 0.022 | 1 (20) | 4 (80) | 0.003 | 1 (20) | 1 (20) | 3 (60) | 0.507 | 5 (100) | 0 (0) | NA |
| 32 (71.1) | 13 (28.9) |  | 36 (80) | 9 (20) |  | 4 (8) | 21 (42) | 25 (50) |  | 0 (0) | 50 (100) |  |
| 34 (69.4) | 15 (30.6) | 0.184 | 37 (75.5) | 12 (24.5) | 0.451 | 7 (12.7) | 21 (38.2) | 27 (49.1) | 0.387 | 3 (6) | 47 (94) | 0.011 |
| 2 (40) | 3 (60) |  | 3 (60) | 2 (40) |  | 0 (0) | 1 (20) | 4 (80) |  | 2 (40) | 3 (60) |  |
| 2 (50) | 2 (50) | 0.378 | 2 (50) | 2 (50) | 0.330 | 1 (25) | 1 (25) | 2 (50) | 0.811 | 2 (50) | 2 (50) | 0.009 |
| 11 (57.9) | 8 (42.1) |  | 13 (68.4) | 6 (31.6) |  | 2 (9.1) | 7 (31.8) | 13 (59.1) |  | 2 (10) | 18 (90) |  |
| 23 (74.2) | 8 (25.8) |  | 25 (80.6) | 6 (19.4) |  | 4 (11.8) | 14 (41.2) | 16 (47.1) |  | 1 (3.2) | 30 (96.8) |  |
| 15 (68.2) | 7 (31.8) | 0.845 | 16 (72.7) | 6 (27.3) | 0.851 | 5 (18.5) | 8 (29.6) | 14 (51.9) | 0.267 | 3 (13.6) | 19 (86.4) | 0.338 |
| 21 (65.6) | 11 (34.4) |  | 24 (75) | 8 (25) |  | 2 (6.1) | 14 (42.4) | 17 (51.5) |  | 2 (6.1) | 31 (93.9) |  |

| MMR |  |  | PD-L1 |  |  |  |  | PD-1 |  |
| --- | --- | --- | --- | --- | --- | --- | --- | --- | --- |
| Intact | Loss | p value | Negative | Low | High | p value | Low | High | p value |
| *n* (%) | *n* (%) |  | *n* (%) | *n* (%) | *n* (%) |  | *n* (%) | *n* (%) |  |
| 33 (91.7) | 3 (8.3) | 1.000 | 1 (2.8) | 12 (33.3) | 23 (63.9) | 0.208 | 12 (33.3) | 24 (66.7) | 0.026 |
| 22 (91.7) | 2 (8.3) |  | 3 (12.5) | 10 (41.7) | 11 (45.8) |  | 15 (62.5) | 9 (37.5) |  |
| 6 (100) | 0 (0) | 0.436 | 0 (0) | 1 (16.7) | 5 (83.3) | 0.366 | 1 (16.7) | 5 (83.3) | 0.141 |
| 49 (90.7) | 5 (9.3) |  | 4 (7.4) | 21 (38.9) | 29 (53.7) |  | 26 (48.1) | 28 (51.9) |  |
| 40 (90.9) | 4 (9.1) | 0.725 | 3 (6.8) | 18 (40.9) | 23 (52.3) | 0.502 | 17 (38.6) | 27 (61.4) | 0.1 |
| 15 (93.8) | 1 (6.3) |  | 1 (6.3) | 4 (25) | 11 (68.8) |  | 10 (62.5) | 6 (37.5) |  |
| 34 (94.4) | 2 (5.6) | 0.184 | 2 (5.6) | 11 (30.6) | 23 (63.9) | 0.378 | 15 (41.7) | 21 (58.3) | 0.845 |
| 15 (83.3) | 3 (16.7) |  | 2 (11.1) | 8 (44.4) | 8 (44.4) |  | 7 (38.9) | 11 (61.1) |  |
| 37 (92.5) | 3 (7.5) | 0.451 | 2 (5) | 13 (32.5) | 25 (62.5) | 0.330 | 16 (40) | 24 (60) | 0.851 |
| 12 (85.7) | 2 (14.3) |  | 2 (14.3) | 6 (42.9) | 6 (42.9) |  | 6 (42.9) | 8 (57.1) |  |
| 7 (100) | 0 (0) | 0.387 | 1 (14.3) | 2 (28.6) | 4 (57.1) | 0.811 | 5 (71.4) | 2 (28.6) | 0.267 |
| 21 (95.5) | 1 (4.5) |  | 1 (4.5) | 7 (31.8) | 14 (63.6) |  | 8 (36.4) | 14 (63.6) |  |
| 27 (87.1) | 4 (12.9) |  | 2 (6.5) | 13 (41.9) | 16 (51.6) |  | 14 (45.2) | 17 (54.8) |  |
| 3 (60) | 2 (40) | 0.011 | 2 (40) | 2 (40) | 1 (20) | 0.009 | 3 (60) | 2 (40) | 0.338 |
| 47 (94) | 3 (6) |  | 2 (4) | 18 (36) | 30 (60) |  | 19 (38) | 31 (62) |  |
| 55 (100) | 0 (0) | NA | 4 (7.3) | 19 (34.5) | 32 (58.2) | 0.487 | 25 (45.5) | 30 (54.5) | 0.814 |
| 0 (0) | 5 (100) |  | 0 (0) | 3 (60) | 2 (40) |  | 2 (40) | 3 (60) |  |
| 4 (100) | 0 (0) | 0.487 | 4 (100) | 0 (0) | 0 (0) | NA | 2 (50) | 2 (50) | 0.974 |
| 19 (86.4) | 3 (13.6) |  | 0 (0) | 22 (100) | 0 (0) |  | 10 (45.5) | 12 (54.5) |  |
| 32 (94.1) | 2 (5.9) |  | 0 (0) | 0 (0) | 34 (100) |  | 15 (44.1) | 19 (55.9) |  |
| 25 (92.6) | 2 (7.4) | 0.814 | 2 (7.4) | 10 (37) | 15 (55.6) | 0.974 | 27 (100) | 0 (0) | NA |
| 30 (90.9) | 3 (9.1) |  | 2 (6.1) | 12 (36.4) | 19 (57.6) |  | 0 (0) | 33 (100) |  |

¶Followup data were available for 54 patients.
§Tumor tissue of only 55 patients could be analyzed.

Single HPV infection was present in 29/40 patients, more common in younger women (62.1%) as compared to mixed infections. Mixed HPV infections were present in 11/40 patients. As compared to patients with single HPV infection those with mixed infection had a higher recurrence (54.5% vs. 29.2%) and death (45.5% vs. 20.8%). Mixed HPV infections with 16/18 and non-16/18 HPV genotypes were present in 3 patients; all of these patients had a recurrence and died subsequently. Mixed non-16/18 HPV infections were seen in 8 patients and had a lower recurrence (37%, p=0.018) and death (20%; p=0.003) as compared to the mixed 16/18 and non-16/18 HPV genotypes (Table 3).

**Table 3.**
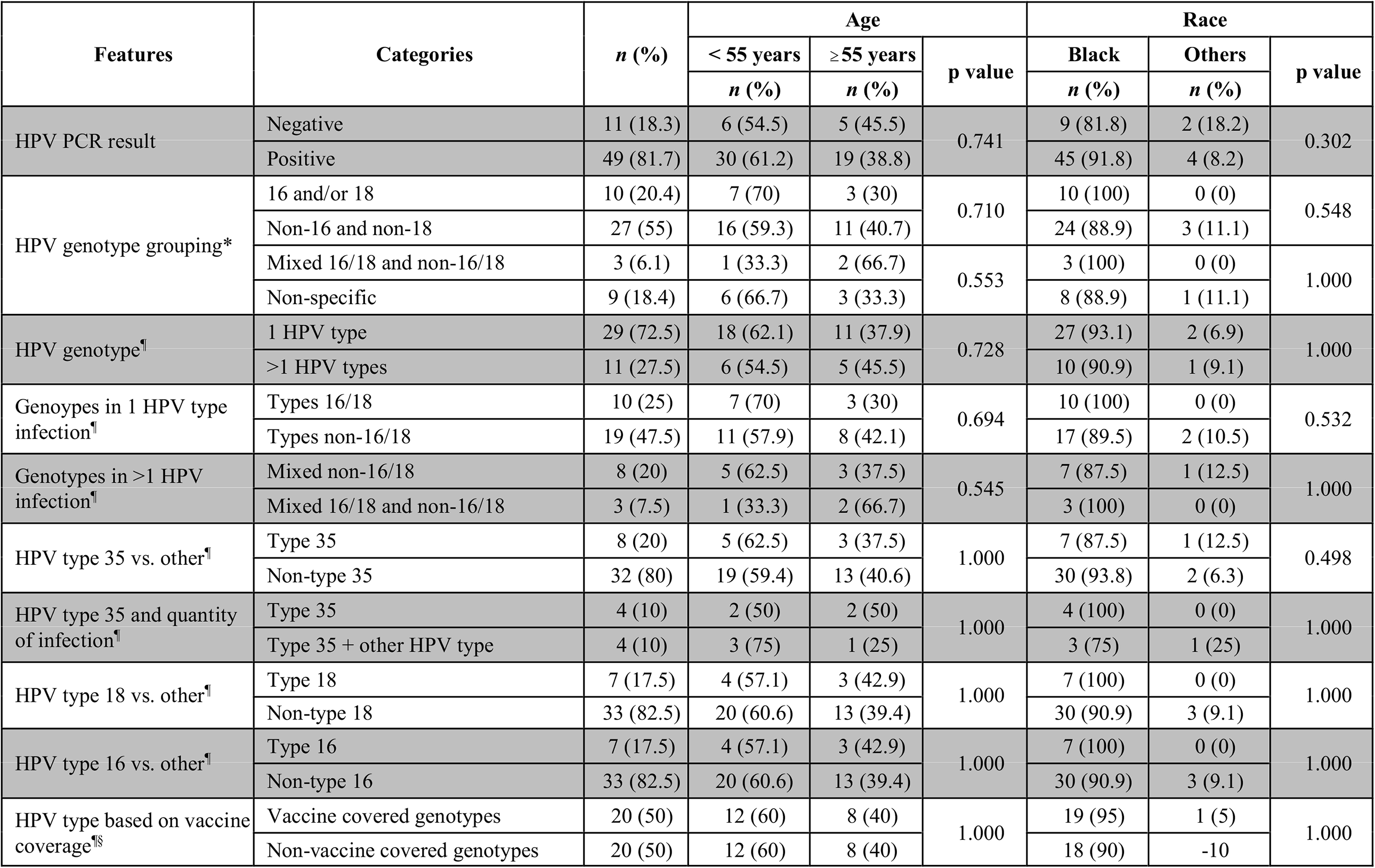
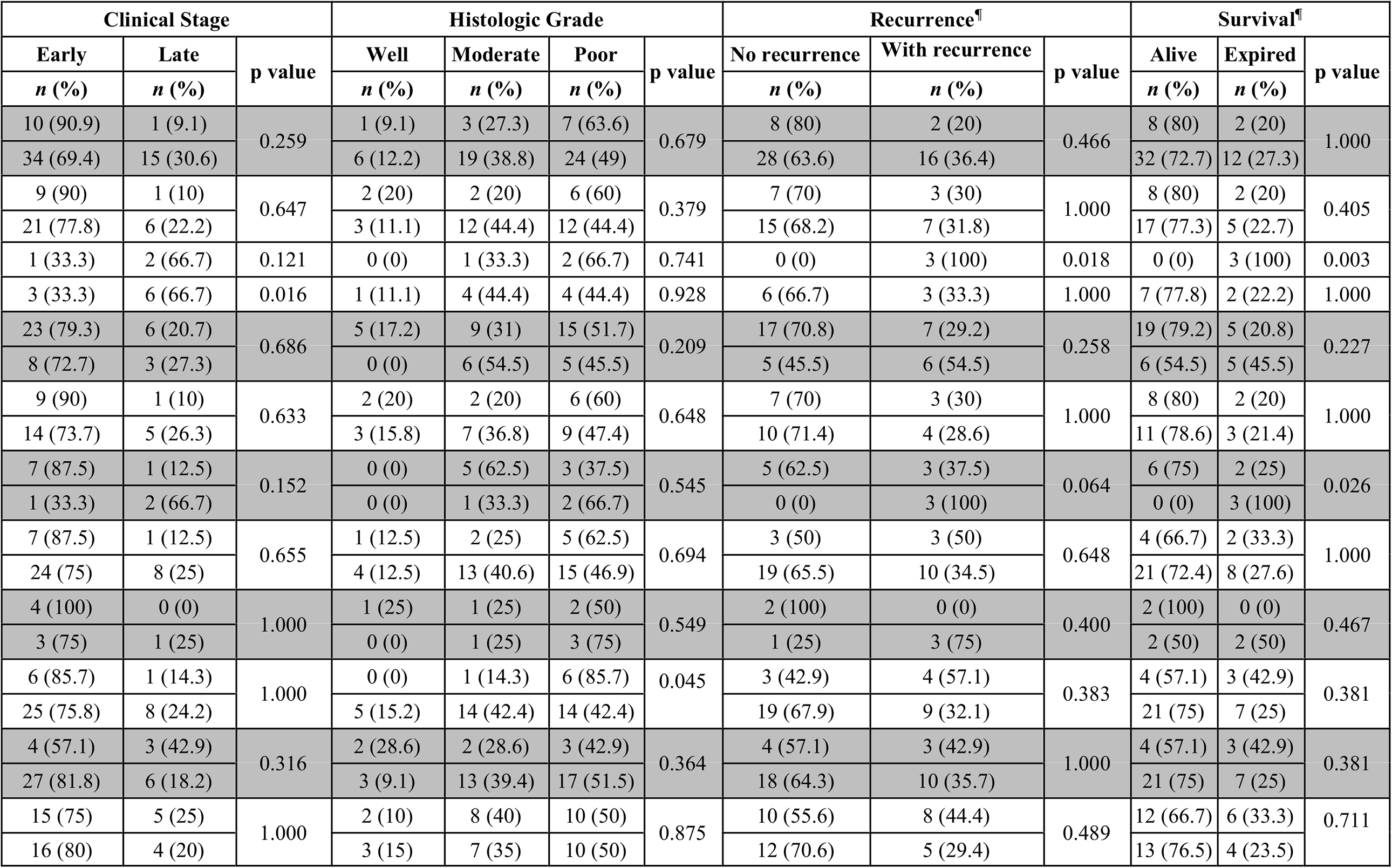
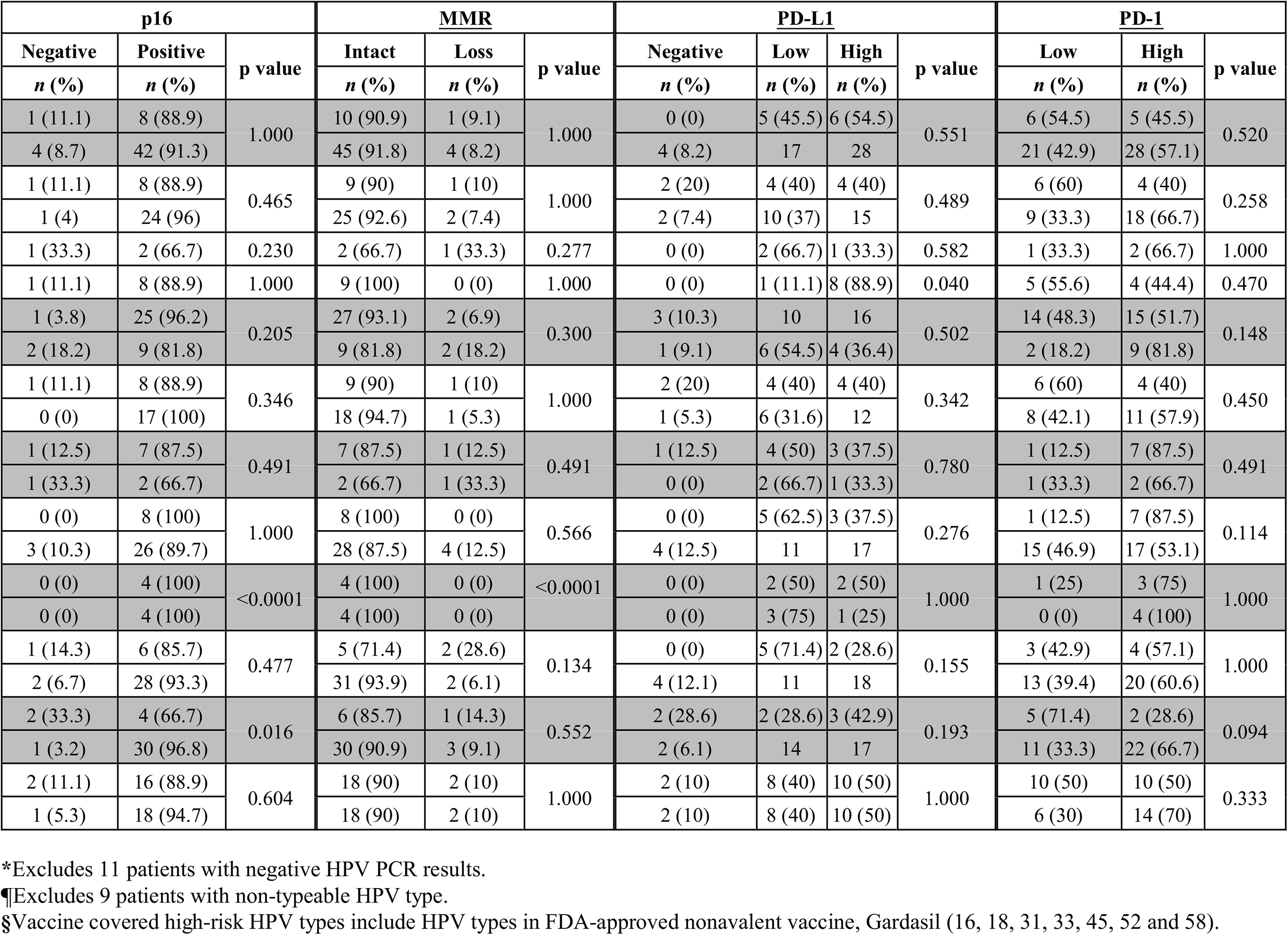
Clinicopathologic characteristics of patients in relation to HPV results.

Patients infected with HPV types covered by the currently available HPV vaccine had a higher recurrence and death (44.4% and 33.3%, respectively) as compared to those with non-vaccine covered HPV (29.4% and 23.5%, respectively).

More patients with negative HPV PCR had poor histologic grade (63.6% vs. 49%) as compared to patients with HPV PCR positive tumors. 2 of 10 (20%) with negative HPV PCR results had recurrence and subsequently died of the disease. There was no significant difference in the prognosis with HPV PCR positive results. 6/9 patients (66.7%) with non-specific HPV PCR had late clinical stage cancer (p=0.016).

5/55 (9.1%) patients lacked p16 expression in the tumor cells, and they were all Black. Loss of p16 correlated with higher recurrence (p=0.022) and death (p=0.003). Patients with negative p16 immunoreactivity showed loss of MMR (40%) (p=0.011) and negative to weak PD-L1 expression (80%; p=0.009), as compared to those with positive p16.

6/60 (10%) patients were Other and had favorable outcomes. All Other patients were diagnosed at an early clinical stage. Only one patient had recurrence and none died during the 60-month follow-up. Most of the Black patients had poor histologic tumor grade at diagnosis (28/55; 51.9%). More Black patients were diagnosed at late stage (16/55, 29.6%), and about a third of them (17/55; 35.4%) had recurrence, 14 of whom died subsequently (14/55; 29.2%).Younger women tend to have early stage cancer (80.6%) at diagnosis as compared to older patients (62.5%). At 5-year follow-up, the mortality rate among older patients (6/21; 28.6%) was slightly higher than that observed among younger patients (8/33; 24.2%).

Recurrence was more frequently observed in patients who presented at a late clinical stage (p=0.028); more than half of the patients (57.1%) diagnosed in late clinical stage and only a quarter (25%) of patients in early clinical stage had recurrence. Similarly, greater number of patients diagnosed at a late clinical stage (42.9%) died as compared to those diagnosed at an early stage (20%). Patients with poor histologic grade tumors had higher recurrence (p=0.022) and death (p=0.046). No recurrence or death was noted in patients with well-differentiated tumors. Most of those patients who had recurrence (77.8%) subsequently died (p<0.0001) (Table 2).

PD-1 expression on TILs was higher in younger patients (66.7%) than those >55 years (37.5%) (p=0.026). It was higher in patients with mixed HPV infections (81.8%) than single infection (54.5) and specifically more in HPV 35 (75%) and other HPV non-16/18 types (66.7%) than HPV 16 (28.6%) and HPV 18 (57.1%). PD-L1 expression was higher on tumor cells in non-black patients (83.3%) as compared to black patients (53.7%). MMR expression was lost in 2/11(18.2%) patients with mixed HPV infections as compared to those with single HPV type (2/29, 6.9%,), but the difference is not statistically significant (p=0.300). There was no correlation between PD-1 and PD-L1 and MMR protein expression. PD-L1 and MMR did not correlate with age, clinical stage or histologic grade (Table 2).

### Survival Analysis

Sixty months after diagnosis, 15 patients were alive and 14 had died of disease; 6 were lost to follow-up soon after the procedure, and 25 had variable follow-up between 1 and 59 months. Eighteen of 54 patients had a recurrence. Patients diagnosed at early and late clinical stages had mean disease-free survival (DFS) of 46.1 and 25.0 months (log rank test p=0.009) and mean overall survival (OS) of 49.99 and 35.78 months, respectively (log rank test p=0.049).

PD-L1 expression showed a positive correlation with DFS and OS. Negative, low and high expression of PD-L1 were associate with mean DFS of 6.1, 27.0 and 36.5 months, and mean OS of 24.9, 28.1 and 40.3 months, respectively. In early clinical stage, high PD-L1 expression was associated with longer DFS (log rank test p=0.006) and OS (log rank test p=0.025)(Figs. 5A, 5B).

**Figure 5.**
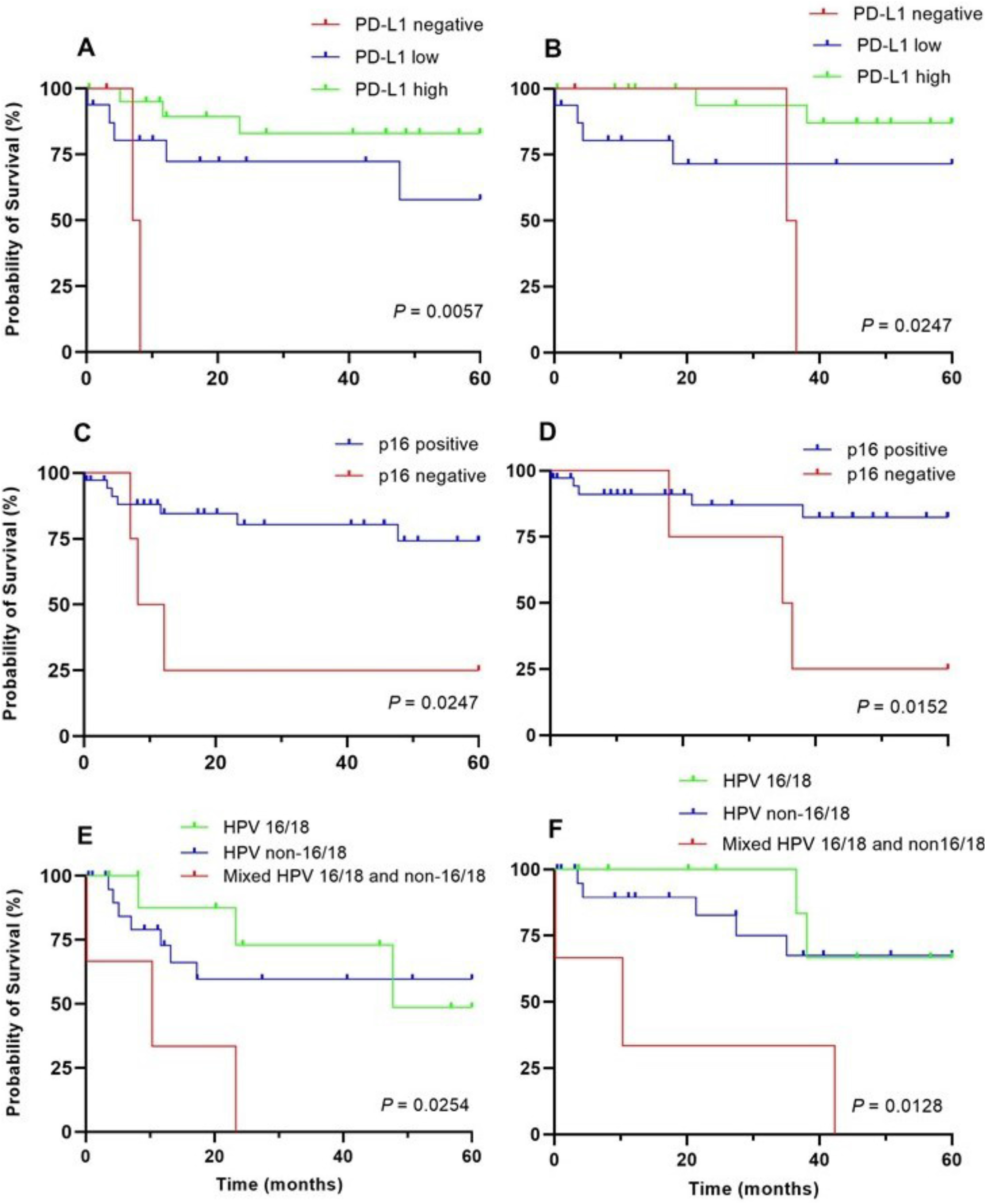
Five-year disease-free (DFS) and overall survival (OS) of patients with cervical cancer diagnosed at early clinical stage. A: DFS based on negative, low and high PD-L1 expression. B: OS based on negative, low and high PD-L1 expression. C: DFS based on positive and negative p16 expression. D: OS based on positive and negative p16 expression. E: DFS based on HPV types. F: OS based on HPV types.

Mean DFS and OS for patients with negative and positive p16 expression was 21.8 and 48.6 months and 7.1 and 12.2 months, respectively. In patients diagnosed at early clinical stage, p16 overexpression on tumor cells was associated with longer DFS (log rank test p=0.033) and OS (p=0.019) (Figs. 5C, 5D).

Patients with non-16/18 infections had shorter mean DFS (24.3 months) and OS (30.1 months) than those with HPV 16/18 (29.8 and 35.3 months) (log rank test p=0.779 and 0.609, respectively). Patients with mixed HPV infections had a shorter mean DFS and OS (18.9 and 25.4 months, respectively, log rank test p<0.0001) as compared to those with single HPV type (26.9 and 32.4 months, respectively), and in patients with negative HPV by PCR (36.6 and 37.2 months, respectively) (Figs. 5E, 5F). Patients with mixed HPV 16/18 and non-16/18 types had even shorter mean DFS and OS (11.3 and 17.6 months, respectively).

Patients with HPV genotype 35 had shorter mean DFS (33.2 months) and OS (18.6 months) compared to those infected with other types in early clinical stage (log rank test p=0.296 and 0.123, respectively). In late clinical stage, however, patients with HPV genotype 18 had significantly shorter DFS and OS (log rank test p=0.025).

When all other variables are controlled using Cox regression analysis, high PD- L1 expression (p=0.003), MMR retention (p=0.01), clinical stage (p=0.048), histologic grade (p=0.015) and mixed HPV infection (p=0.026) remained independent predictors of DFS.

## Discussion

The results of this study confirm reported higher prevalence of non-16/18 HPV genotypes among Black women with cervical cancer. However, in contrast to published data, our patients with non-16/18 HPV types had worse prognosis. Those with mixed HPV type infection had even poorer outcome. Also, patients who tested negative for HPV by PCR and negative for p16 on immunohistochemistry (IHC) had poor survival. We also found high rate of PD- L1 expression in cervical cancer cells, which was surprisingly associated with better survival. Our findings suggest a unique tumor microenvironment in squamous cell carcinoma in Black women which could offer therapeutic opportunities leading to survival benefit.

Of the 120 known HPV genotypes, about 40 infect the human genital mucosa and 15 have been classified as high-risk types (16, 18, 31, 33, 35, 39, 45, 51, 52, 56, 58, 59, 68, 73, and 82) ^19-21^. In our predominantly Black population, we identified 13 of these high-risk HPV types of which type 35 was most frequent (20.0%) followed by types 16 and 18 (each 17.5%). HPV non-16/18 (75%) were more frequent than HPV 16/18 (25%) types. Distribution of 16/18 and non- 16/18 has been found to be variable in different parts of the world. In the US a recent population-based study of 693 patients with invasive cervical carcinoma showed a predominance of HPV 16 (51.4%) followed by non-16/18 (24.2%) and HPV 18 (15.9%), while 8.5% patients were negative for HPV ^21^.

Outside of the US a higher prevalence of non-16/18 HPV high-risk types have been reported than HPV 16/18 genotypes, and the former were found to be associated with a higher grade of cervical disease ^22^. In Korea HPV 16 and 58 were found to be in almost equal frequency in a cervical cytology study of 1158 women; patients with HPV 58 had a significantly higher progression rate to high grade dysplasia ^23^.

A meta-analysis of HPV types in invasive cervical cancer in Asia found that types 16, 18, 52 and 58 had similar prevalence ^24^. Studies from the Middle East have also shown higher prevalence of non-16/18 HPV genotypes, with HPV 31 most prevalent in Yemen and HPV 33 in Kuwait followed by 35, 39, 45, 52 and 58 ^25,26^. These studies indicate that the prevalence of non-16/18 HPV types differs globally and their impact in different patient populations has been largely under-estimated.

Studies on HPV vaccine effectiveness have highlighted protection against HPV-related pre-cancers and cancers, with a recent meta-analysis estimating 51% reduction in high grade cervical dysplasia among 15-19-year-olds and 31% reduction among 20-24-year-olds ^27-28^. In light of these encouraging results, the findings of our study are striking because the currently available 9-valent HPV vaccine (against types 6, 11, 16, 18, 31, 33, 45, 52, 58) does not include seven HPV types (35, 39, 51, 53, 56, 59, 68) that were detected in about one-third (33.3%) of our patients, mostly of Caribbean origin. Vaccines currently available to prevent the most common strains of HPV, may not offer the same level of protection in our Black population and other racial and ethnic groups globally. The diversity of patients and the prevalence of different HPV genotypes in the US and globally should be acknowledged as scientists guide further HPV vaccine development. To ensure successful cervical cancer prevention, physicians should continue to perform cervical cancer screening in women until complete high-risk HPV virus coverage is available in the vaccine.

Our patients with non-16/18 types had worse prognosis than those infected with 16/18 in early clinical stage. This finding fails to validate earlier studies which found that women with HPV 16/18 positive tumors had worse survival than those with other HPV types ^21,29,30^.

One fifth of our patients had a mixed HPV infection and theywere all found to have higher grade carcinomas (moderately to poorly differentiated) and a lower DFS and OS than those with a single HPV type infection. These tumors also had a higher PD-1 expression in TILs making them high-risk for aggressive course of disease although they could also prove to be targets for checkpoint inhibitor therapy.

Our patients with negative HPV PCR results and non-typeable HPV types had high tumor grade at diagnosis and were also diagnosed at a late clinical stage. These findings suggest that patients with non-typeable oncogenic HPV types should be treated with higher circumspection because of their aggressive behavior and limited knowledge about their genotype making them hard to include in the vaccines. Patients with negative p16 expression had significantly shorter DFS and OS as compared to those with p16-positive tumors. Although this observation is based on a small sample in our study, similar observations in other population studies have recorded poor survival among women negative for both HPV PCR and p16 expression ^21,31-35^. Only 9% of our patients had negative p16 expression, a finding similar to those reported in other studies on patients with invasive squamous cell carcinoma of the cervix. Among our patients with negative HPV PCR results, 8 of 9 had positive p16 expression. This contradictory pattern has been described in other studies which found the initial HPV PCR negativity – with p16 positive IHC – reduced from 19/209 to 5/209 cervical carcinomas after expanding molecular studies (targeting E6/E7 genes) ^36^. The p16 positivity is therefore considered a more sensitive indicator of HPV-mediated carcinogenesis in cervical cancers. Loss of the frequently targeted viral L1 gene after viral integration could be a reason for HPV-negative test results,^37^ while other studies have identified methodological limitations (long-term storage, low tumor tissue) as probable causes ^38^.

PD-L1 expression in cervical cancer in our patients was 93% which far exceeds the reported frequency of 35-70% ^9-11^. Expression of PD-L1 has been reported to be of variable prognostic information in neoplasms of different organs. It is a poor prognostic indicator in solid tumors, including esophageal, gastric, colorectal, and pulmonary adenocarcinoma, and a good prognostic indicator in lung squamous cell carcinoma ^39,40^. Our study found PD-L1 to be a predictor of longer survival in early stage cervical carcinoma. Presence of higher PD-L1 increases the possibility of using targeted immunotherapy in these patients.Racial difference in PD-L1 expression of tumor cells had not been widely investigated. A recent single institution study of 114 cases, 36% of which were Black, concluded that PD-L1 expression in non-small cell lung cancer did not differ by race, clinical stage or smoking history ^41^. Findings of a higher PD-L1 expression on cervical carcinoma cells in our population should be explored further in a larger multiracial study to identify the association of race with PD- L1 expression.

PD-1 expression in TILs surrounding cervical carcinoma showed no association with clinical stage, histologic grade, or HPV type, although PD-1 expressing TILs were greater in younger patients and patients with mixed and non-16/18 HPV infections. The prognostic significance of PD-1 expression in cervical carcinoma is still not fully elucidated, however, increase in PD-1 expression has been associated with poor prognosis in gastric, renal, nasopharyngeal and non-small cell lung carcinomas,^42-45^ and increased survival in melanoma, glioblastoma, and ovarian, breast, and primary HPV-positive head and neck cancers ^46-50^. Although not of prognostic value in our study, higher expression of PD-1 may be helpful to guide immunotherapy in the treatment of cervical carcinoma in patients with HPV infection of more severe types like non-16/18 and mixed, and patients with high stage carcinoma with limited treatment options.

MMR deficiency in solid tumors has recently been linked to susceptibility to immunotherapies targeting the PD-1/PD-L1 axis ^14,15^. Loss of MMR proteins has been shown to correlate with tumoral PD-L1 expression in colorectal and endometrial carcinomas, but this association has not been examined in cervical carcinoma, where MMR deficiency is less common. In the current study 8.3% (5/60) patients were MMR-deficient whereas other reports have documented 25.8% (17/66) cervical carcinoma patients with MMR deficiency and significant correlation with PD-L1 expression on tumor cells ^51^. All of our five MMR-deficient patients expressed PD-L1.

Race, specifically the Black race, has been identified as one of the most common demographic factors associated with cervical squamous cell carcinomas ^21^. Blacks have worse survival rates likely due to more advanced-stage tumors at diagnosis and less opportunity likely to receive adequate treatment as compared with white women ^52-54^. This trend was similarly reflected in our study where more than half of the Black patients had poor histologic tumor grade at diagnosis, about a third were diagnosed at late clinical stage and a third had recurrence who subsequently died. On the other hand, all non-Black patients, albeit in small number, had favorable prognosis; all of them were diagnosed at an early stage and all survived.

## Conclusions

Most Black patients in our study were infected with non-16/18 HPV genotypes, which were associated with a lower disease-free and overall survival. HPV 35 was the most common type isolated in our patient population, and this HPV genotype is not included in the commercially available nonavalent vaccine. Exclusion of HPV 35 and other common genotypes (39, 51, 53, 56, 59, 68) from the vaccine exacerbates mortality from cervical cancer in these underserved communities. Patients with HPV 35 and those with mixed HPV infection had shorter DFS and OS, especially in early clinical stages. A remarkably high percentage of PD-L1 expression in cervical cancer cells was observed in our predominantly Black patient population and a high PD-1 expression on TILs in younger patients. These findings suggest that immune checkpoint inhibitors could be given consideration in the management of primary invasive squamous cell carcinoma of the cervix among Black patients.

## Data Availability

Available upon request

## Acknowledgments

The study was approved by the Institutional Review Board & Privacy Board (IRB) of the State University of New York, Downstate Health Sciences University (IRB #1230970-3). We would like to express our gratitude to Ms. Mengru Li for her expert assistance in immunophenotyping of tumors for PD-L1 (SP263), PD- 1, MMR proteins (MSH2, MSH6, PMS2 and MLH1) and p16 expression.

## Author Contributions

RPM, YYZ, WCL, YCL, RG designed the research study. YYZ, WCL and YCL acquired the case list and clinical information. YYZ recorded clinical follow up. RPM, RW, EG, TH and CC conducted the experiments and acquired laboratory data. RW and RG provided reagents and laboratory space. MAH prepared the figures. RPM, RG, ZB and MAH analyzed the data and wrote the manuscript. All authors reviewed the manuscript.

## Supplementary Information

**Supplementary Table.**
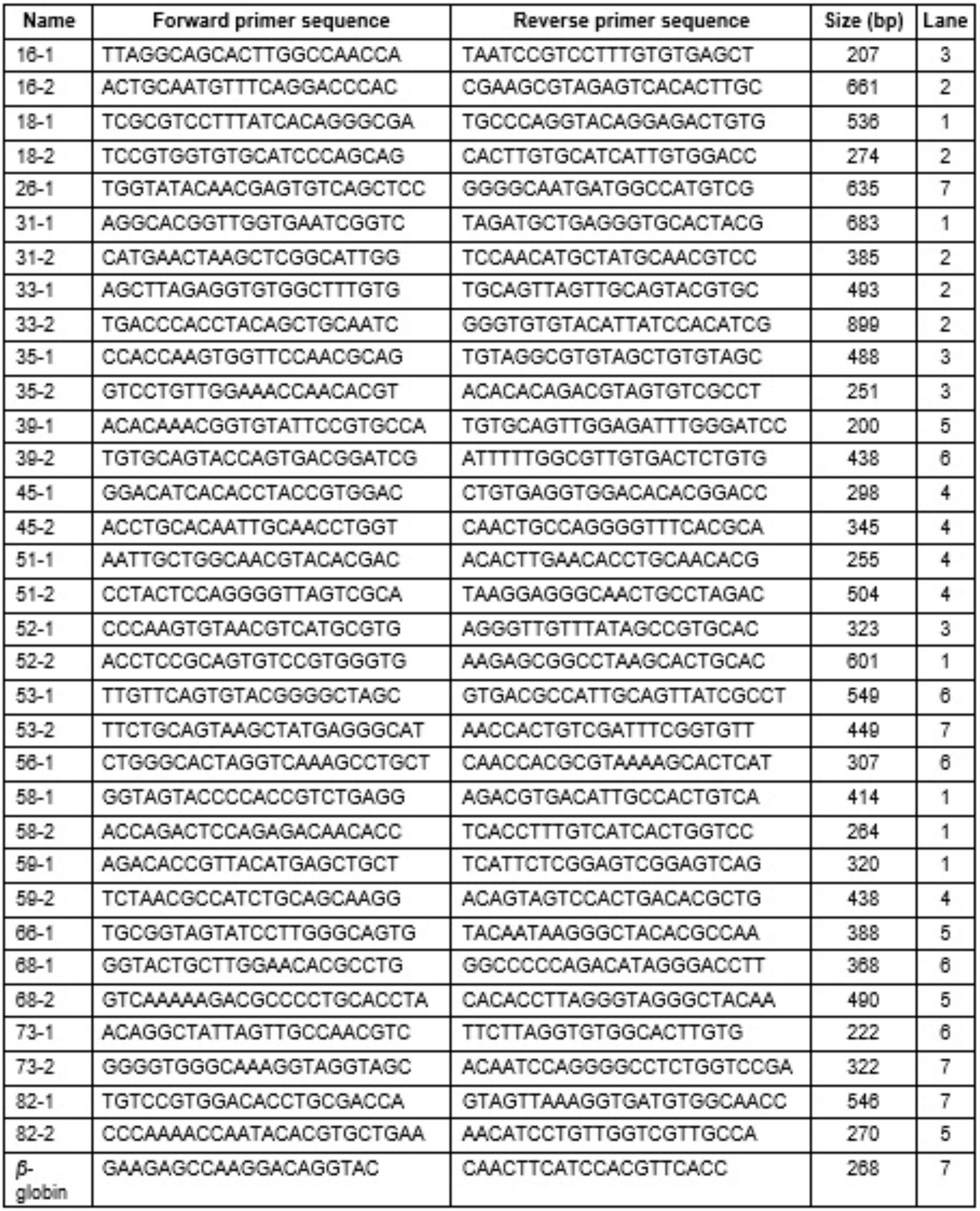
Primers used for the PCR analysis of high-risk HPV genotypes.

